# Can electroencephalography (EEG) identify ADHD subtypes? A systematic review

**DOI:** 10.1101/2022.03.25.22272900

**Authors:** Jessica Slater, Ridha Joober, Brenda Lynn Koborsy, Samantha Mitchell, Ella Sahlas, Caroline Palmer

## Abstract

Attention Deficit/Hyperactivity Disorder (ADHD) has been associated with atypical patterns of neural activity measured by electroencephalography (EEG). However, the identification of EEG diagnostic biomarkers has been complicated by the disorder’s heterogeneity. The objective of this review was to synthesize the literature investigating EEG variation in patients diagnosed with ADHD, addressing the following questions: 1) Are the diagnostic ADHD subtypes associated with different EEG characteristics? 2) Are EEG measures correlated with ADHD traits and/or symptom severity? and 3) Do classification techniques using EEG measures reveal different clinical presentations of ADHD? Outcomes highlight the potential for electrophysiological measures to provide meaningful insights into the heterogeneity of ADHD, although direct translation of EEG biomarkers for diagnostic purposes is not yet supported. Key measures that show promise for the discrimination of existing ADHD subtypes and symptomatology include: resting state and task-related modulation of alpha, beta and theta power, and the event-related N2 and P3 components. Prescriptions are discussed for future studies that may help to bridge the gap between research and clinical application.

## 1. Introduction

ADHD is the most common neurodevelopmental disorder (Polanczyk et al., 2007; Simon et al., 2009), yet the search for definitive biomarkers has met with limited success; diagnosis of ADHD relies primarily on self-report questionnaires and clinical interviews, and treatment often takes the form of a trial-and-error pharmacological approach. Although abnormalities in neural activation have been associated with ADHD, several reviews evaluating the utility of electroencephalography (EEG) have concluded that it is not reliable enough for clinical diagnosis in either children or adults (Adamou et al., 2020; Arns et al., 2013; Lenartowicz and Loo, 2014). A clearer understanding of the relationship between EEG characteristics and ADHD symptom profiles could lead to the development of more objective diagnostic tools as well as to help guide individualized treatment strategies.

In this systematic review, we assess the heterogeneity of ADHD by addressing three distinct aims: 1) Are the diagnostic ADHD subtypes associated with different EEG characteristics? 2) Are EEG measures correlated with ADHD traits and/or symptom severity? and 3) Do classification techniques using EEG measures reveal different clinical presentations of ADHD? The diagnostic criteria for ADHD include the presence of either a) frequent inattentive symptoms; b) frequent hyperactive/impulsive symptoms; or c) both inattentive and hyperactive/impulsive symptoms, in multiple settings (such as home, school or work; with friends or relatives; in other activities). According to the DSM-5, children must present with six or more symptoms in either of these two categories; adults must present with five or more symptoms in either category, and symptoms must also have been present during childhood. The three diagnostic “subtypes” (DSM-IV) or “clinical presentations” (DSM-5) indicate the categories in which diagnostic criteria are met: predominantly inattentive, predominantly hyperactive/impulsive, and combined subtype, in which the diagnostic criteria are met in both symptom domains. For clarity, the term “subtype” is used throughout this review to refer to the predominantly inattentive subtype, predominantly hyperactive/impulsive subtype, and combined subtype classifications. The terms “presentation” and “clinical presentation” are used more generally to refer to the pattern of behavioral symptoms in an individual with ADHD.

The shift in terminology from “subtype” in DSM-IV to “presentation” in DSM-5 was supported by a multi-dimensional approach to understanding ADHD symptoms (Epstein and Loren, 2013), consistent with the RDoC (Research Domain Criteria) measurement system advanced by the NIMH (Insel et al., 2010). Following this view, we assess not only diagnostic subtypes but also the variation in inattentive or hyperactive/impulsive symptoms that do not necessarily meet diagnostic criteria for ADHD subtypes, referred to in this review as symptom dimensions. In addition to studies comparing group differences in clinical subtypes, we have therefore included studies assessing correlational relationships between ADHD symptoms and EEG measures in Aim 2.

Finally, increasing evidence suggests that the multiple neurophysiological mechanisms contributing to ADHD symptoms may result in EEG differences that are not necessarily aligned with the current diagnostic subtypes. Our third aim therefore considers studies in which subgroups of ADHD are identified based on EEG characteristics separate from diagnostic criteria; for example, by using cluster analysis or latent classifier techniques. In combination, these three aims provide a synthesis of current research in which the variation in EEG characteristics is investigated in patients diagnosed with ADHD.

## 2. Methods

### 2.1 Pre-registration and search strategy

The current review was performed in compliance with the PRISMA guidelines for systematic reviews and meta-analyses (Page et al., 2021). The review protocol was submitted to PROSPERO international prospective register of systematic reviews on June 20^th^, 2020. Due to the COVID-19 pandemic, the registration record was automatically published without further review by the PROSPERO team on July 23^rd^, 2020 and is available online at https://www.crd.york.ac.uk/prospero/. Search terms included keywords related to ADHD, EEG, electroencephalography, evoked potentials and ERP, and the search included age ranges from 6 to 44 years. Given advances in EEG methods over the past decade, the search was limited to studies published in or after 2009. The search strategy was peer reviewed by an expert in the field. The searches were carried out on the PubMed, PsycINFO and Web of Science databases on August 12th, 2020. Search results were filtered to include only peer-reviewed primary research articles that examined human subjects and were available in English.

### 2.2 Inclusion criteria

Studies were included if they examined EEG characteristics in patients diagnosed with ADHD, using one or more of the following analyses: 1) Comparison of EEG characteristics between two or more subtypes of ADHD; 2) Examination of relationships between EEG measures and ADHD traits or symptom severity within an ADHD sample, or 3) Identification of two or more EEG-defined subgroups within an ADHD sample. The studies had to include participants with a clinical diagnosis of ADHD, according to DSM-IV or DSM-5 criteria.

### 2.3 Exclusion criteria

Studies were excluded if the ADHD participants had epilepsy or significant psychiatric comorbidities. Studies were excluded if they focused primarily on changes in symptomatology within individuals across the lifespan. Training and intervention studies, e.g. assessing neurofeedback or medication efficacy, were excluded, as were studies focused on assessing the impact of independent risk factors, e.g. genetic factors, birth weight, exercise or general fitness. Studies highlighting new methods were excluded if the study goals were focused on evaluating the method rather than investigating ADHD. Studies combining EEG with other methods (e.g. fNIRS or skin conductance measures) were excluded if the non-EEG methods were necessary for interpretation of the primary research outcomes.

### 2.4 Screening and data extraction

An initial screening of titles and abstracts was conducted by three co-authors; any discrepancies were resolved through discussion. In cases where the abstract described only comparisons between ADHD and control groups but other inclusion criteria were met, the article was flagged for further screening to determine if additional subtype or correlational analyses were carried out. Finally, a full-text review was carried out on the remaining articles to extract the following data, where available: author, year of publication, demographics, ADHD subtypes and sample sizes, experimental tasks and EEG measures (see Tables 1–3, corresponding to Aims 1-3).

**Table 1.** Study characteristics of included studies evaluating EEG characteristics in clinical ADHD subtypes (Aim 1).

| **Author (yr)** | **Age (years)** | **ADHD subtypes and participants** | **Tasks** | **EEG analyses** | **Quality rating / 20** |
| --- | --- | --- | --- | --- | --- |
| Bluschke, 2017 | 7-15 | ADHD-C = 34; ADHD-I = 25 | Visual flanker task | P1, N1, N2, P3 | 18 |
| Bluschke, 2018 | 11 | ADHD-C = 16; ADHD-I = 16; CTRL = 16 | Time interval estimation task | P1, N1, CNV, FRN | 17 |
| Dupuy, 2011 | 8-12 | ADHD-C = 30; ADHD-I = 30; CTRL = 30 (all females) | Resting state EEG, eyes closed | delta, theta, alpha, beta | 13 |
| Dupuy, 2013 | 8-12 | ADHD-C = 80; ADHD-I = 80; CTRL = 80 | Resting state EEG, eyes closed | delta, theta, alpha, beta | 17 |
| Dupuy, 2014 | 7-12 | ADHD-C = 20; ADHD-I = 20; CTRL = 20 | Resting state EEG, eyes closed | delta, theta, alpha, beta | 16 |
| Ghaderi, 2017 | 7-11 | ADHD-C = 12; ADHD-I = 10; CTRL = 13 | Resting state EEG, eyes open | theta, alpha, beta1, beta2, beta3 | 11 |
| Gong, 2014 | 6-14 | ADHD-C = 16; ADHD-I = 15; CTRL = 15 | Gambling task | FRN, LPP | 14 |
| Gonzalez-Castro, 2010 | 6-12 | ADHD-C = 57; ADHD-I = 54; ADHD-H = 53; CTRL = 56 | Resting state EEG | beta, theta | 19 |
| Gorman Bozorgpour, 2013 | 18-50 | ADHD-C = 22; ADHD-I = 18; CTRL = 38 | Go/No-Go task | LRP, P3b | 14 |
| Heinrich, 2014 | 8-11 | ADHD-C = 15; ADHD-I = 9; CTRL = 19 | Attention Network Test (ANT) | P3, theta, alpha, beta, theta | 13 |
| Inoue, 2010 | 9-13 | ADHD-C = 6; ADHD-I = 6; CTRL = 12 | Go/No-Go Continuous Performance Test (CPT) | N1, N2, P3 | 13 |
| Johnstone, 2009 | 8-14 | ADHD-C = 15; ADHD-I = 15; CTRL = 15 | Visual Go/No-Go task | N1, P2, N2, P3, CNV-E, CNV-L | 13 |
| Kratz, 2011 | 8-11 | ADHD-C = 15; ADHD-I = 10; CTRL = 19 | Attention Network Test (ANT) | P3, CNV | 15 |
| Mazaheri, 2014 | 12-17 | ADHD-C = 17; ADHD-I = 17; CTRL = 23 | Cued flanker task | alpha, beta, FRN, LPP | 11 |
| Sanchez-Gonzales, 2017 | 7-14 | ADHD-C = 7; ADHD-I = 6; ADHD-H = 7; CTRL = 23 | Observation-execution task | alpha, mu | 17 |

**Table 2.** Study characteristics and EEG measures of included studies assessing correlations between EEG measures and ADHD traits and/or symptom severity (Aim 2).

| First Author (year) | Age (years) | ADHD subtypes and participants | Tasks | EEG Measures | Quality rating / 20 |
| --- | --- | --- | --- | --- | --- |
| Abramov, 2019 | 11-13 | ADHD = 19; CTRL = 20 | Attention network test (ANT) | P3 | 15 |
| Alperin, 2017 | ADHD: M = 13.70 +/- 1.48; CTRL: M = 13.87 +/- 1.08 | ADHD-C = 16; ADHD-I = 30; ADHD-H = 2; CTRL = 60 | Emotional Go/No-Go task | P1, N170, P3, LPP | 18 |
| Barry, 2010 | 8-12 | ADHD-C = 25; ADHD-I = 15; CTRL = 40 | Resting state EEG, eyes closed | delta, theta, alpha, beta, gamma | 16 |
| Barry, 2011 | 8-13 | ADHD-C = 25; ADHD-I = 15; CTRL = 40 | Resting state EEG, eyes closed | delta, theta, alpha, beta, gamma | 14 |
| Buyck, 2015 | 18-55 | ADHD-C = 15; ADHD-I = 9; CTRL = 20 | 2-choice RT task and flanker task | theta, alpha, beta | 16 |
| Cheung, 2017 | ADHD: M = 18.28 +/- 2.98; CTRL: M = 17.76 +/- 2.16 | ADHD-C = 93; CTRL = 174 | 4-choice RT task | P3, CNV | 15 |
| Cross-Villasana, 2015 | ADHD: 17-48; CTRL: 19-48 | ADHD = 15; CTRL = 15 | Visual search task | PCN | 14 |
| Czobor, 2017 | ADHD: M = 39.6 +/- 9.7; CTRL: M = 30.1 +/- 9.0 | ADHD-C = 22; CTRL = 29 | Go/No-Go task | ERN, Pe | 16 |
| Ellis, 2017 | 7-14 | ADHD = 25; CTRL = 25 | Go/No-Go task | alpha | 17 |
| Grane, 2016 | ADHD: 19-51; CTRL: 18-51 | ADHD-C = 33, CTRL = 31 | Visual Go/No-Go task | P3, CNV, N2 | 17 |
| Guo, 2019 | 8-13 | ADHD = 21; CTRL = 24 | Covert attention task | alpha | 16 |
| Helps, 2010 | 13-16 | ADHD-C = 16; CTRL = 16 | Resting state EEG, eyes open; 2-choice RT task | delta | 16 |
| Ibanez, 2011 | ADHD: M = 33.1 +/- 3.6; CTRL: M = 33.0 +/- 3.8 | ADHD-C = 8; ADHD-I = 2; CTRL = 10 | Dual valence task | N170 | 14 |
| Janssen, 2015 | 7-14 | ADHD-C = 28; ADHD-I = 8; CTRL = 49 | Auditory oddball task | P3b | 17 |
| Kakuszi, 2016 | ADHD: M = 31.6<br>+/- 12.1; CTRL: M = 32.9 +/- 12.8 | ADHD-C = 33; CTRL = 29 | Go/no-go task | RPNS | 17 |
| Kaur, 2019 | ADHD: M = 20.3<br>+/- 1.12; CTRL: M = 20.6 +/- 1.28 | ADHD = 35; CTRL = 35 | Visual Continuous Performance Task (CPT) | N1, N2, P3 | 16 |
| Kochel, 2012 | 19-33 | ADHD = 15; CTRL = 15 (all males) | Emotional Go/No-Go task | LPP | 13 |
| Koehler, 2009 | 18-55 | ADHD-C = 34; CTRL = 34 | Resting state EEG, eyes closed | delta, theta, alpha, beta | 14 |
| Lee, 2020 | 6-18 | ADHD = 16; subclinical ADHD = 13 | Auditory oddball task | MMN | 17 |
| Micoulaud-Franchi, 2019 | ADHD: M = 30.25<br>+/- 7.92 | ADHD = 24 | Conditioning-testing and oddball tasks | P50, P300 | 15 |
| Ogrim, 2012 | 7-16 | ADHD-C = 42; ADHD-I = 20; CTRL = 39 | Resting state EEG, eyes open | theta, beta | 16 |
| Roh, 2015 | ADHD: M = 9.8<br>+/- 1.61; CTRL: M = 9.4 +/- 0.88 | ADHD = 15; CTRL = 20 | Resting state EEG, eyes closed | delta, theta, beta, gamma | 16 |
| Shushakova, 2018 | ADHD: M = 31.21<br>+/- 8.27; CTRL: M = 31.08 +/- 8.83 | ADHD-C = 21; ADHD-I = 18; CTRL = 40 | Visual image task and emotion regulation task | LPP | 17 |
| Shushakova, 2018 | ADHD: M = 31.15<br>+/- 8.24; CTRL: M = 30.59 +/- 8.95 | ADHD-C = 22; ADHD-I = 17; CTRL = 41 | Verbal dot-probe task | N2pc, P1 | 17 |
| Thoma, 2015 | 20-49 | ADHD-C = 14; CTRL = 14 | Reward-based learning task | N100, P200, FRN, P300 | 16 |
| Vollebregt, 2015 | 8-15 | ADHD = 9; CTRL=17 | Resting state EEG, eyes closed | theta, beta | 13 |
| Woltering, 2013 | ADHD: M = 25.1<br>+/- 5.2; CTRL: M = 25.2 +/- 5.2 | ADHD = 54; CTRL = 29 | Go/No-Go task | N2, P3, P1, N1 | 16 |
| Zhang, 2019 | 8-15 | ADHD-C = 21; ADHD-I = 32; CTRL = 37 | Resting state EEG, eyes closed | delta, theta, alpha, beta | 16 |

**Table 3.**
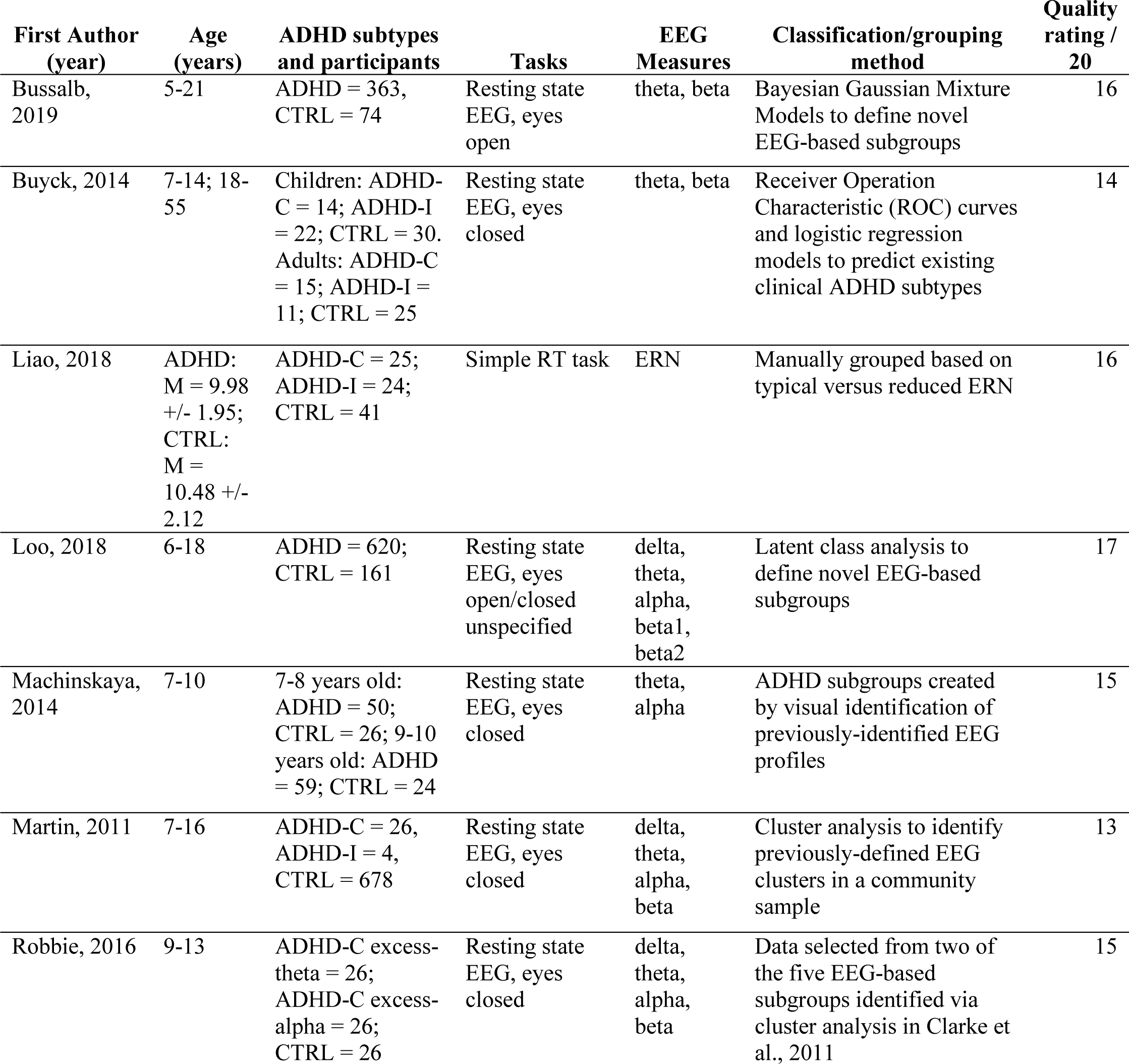
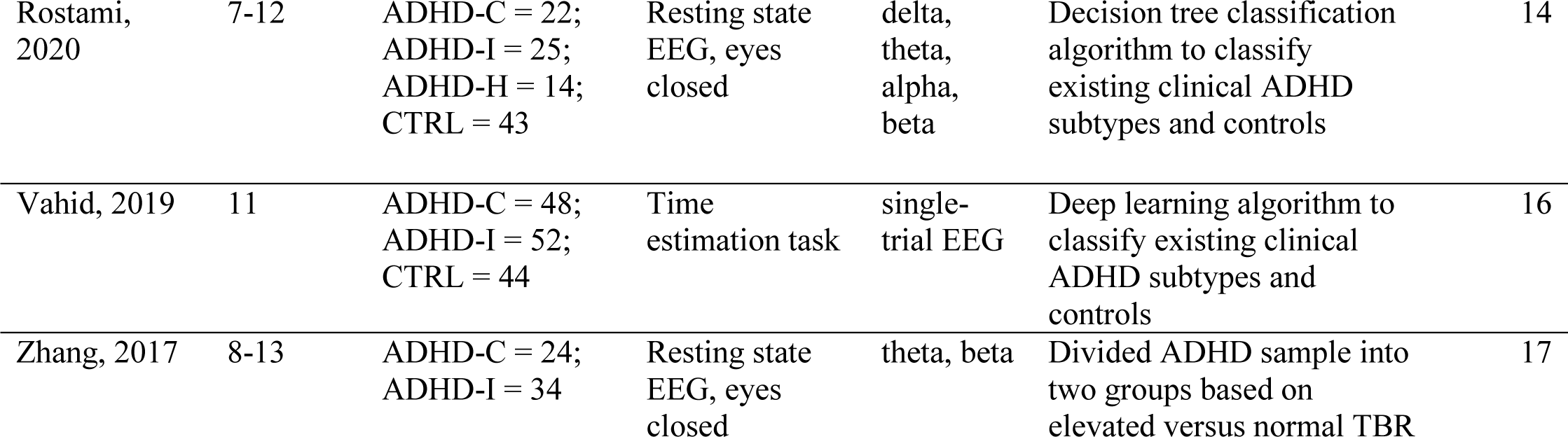
Study characteristics, EEG measures and classification methods of included studies assessing the use of classification techniques with EEG measures to reveal different clinical presentations of ADHD (Aim 3).

### 2.5 Quality assessment

Study quality was assessed using a modified version of the Downs and Black evaluation tool (Downs and Black, 1998), which provides a quality rating checklist for assessing randomized and nonrandomized studies. The checklist evaluates the following areas: (i) reporting (ii) external validity, (iii) bias, (iv) confounding variables, and (v) power. The tool was modified in order to apply to the type of studies included in the review, following a similar approach to that used in a recently published systematic review on a related topic (the use of EEG in diagnosis of adult ADHD) (Adamou et al., 2020). With these modifications, the maximum possible score was 20 and the minimum possible score was zero. Initial ratings for each study were provided by SM, BK and ES (two raters per study); scores were finalized through group discussion with JS. Scores of 17-20 were classified as excellent, 14–16 as good, 11–13 as fair, and <11 as poor. Study quality scores are listed in the summary tables for each aim. (Tables 1–3).

## 3. Results

The searches retrieved 2402 articles across the three databases; once duplicates were removed, 1526 potentially eligible manuscripts remained. The initial screening of titles and abstracts identified 132 articles for further full-text evaluation. In total, 53 studies met the inclusion criteria and were included in the review. Each study aligned with one of the three aims. Figure 1 presents the number of studies identified at each stage of the screening process in a flow diagram. Figure 2 presents the proportion of total studies that examined the three diagnostic subtypes of ADHD (predominantly inattentive, ADHD-I; predominantly hyperactive/impulsive, ADHD-H; and combined, ADHD-C) in their analyses. The largest proportion of studies compared ADHD-I with ADHD-C. The smallest proportion of studies compared all three subtypes, indicating the need for further research. Figure 3 presents the proportion of total studies by the experimental tasks used during EEG recording. The greatest proportion of studies measured resting state EEG and/or inhibitory control tasks; the smallest proportion measured timing tasks.

**Figure 1.**
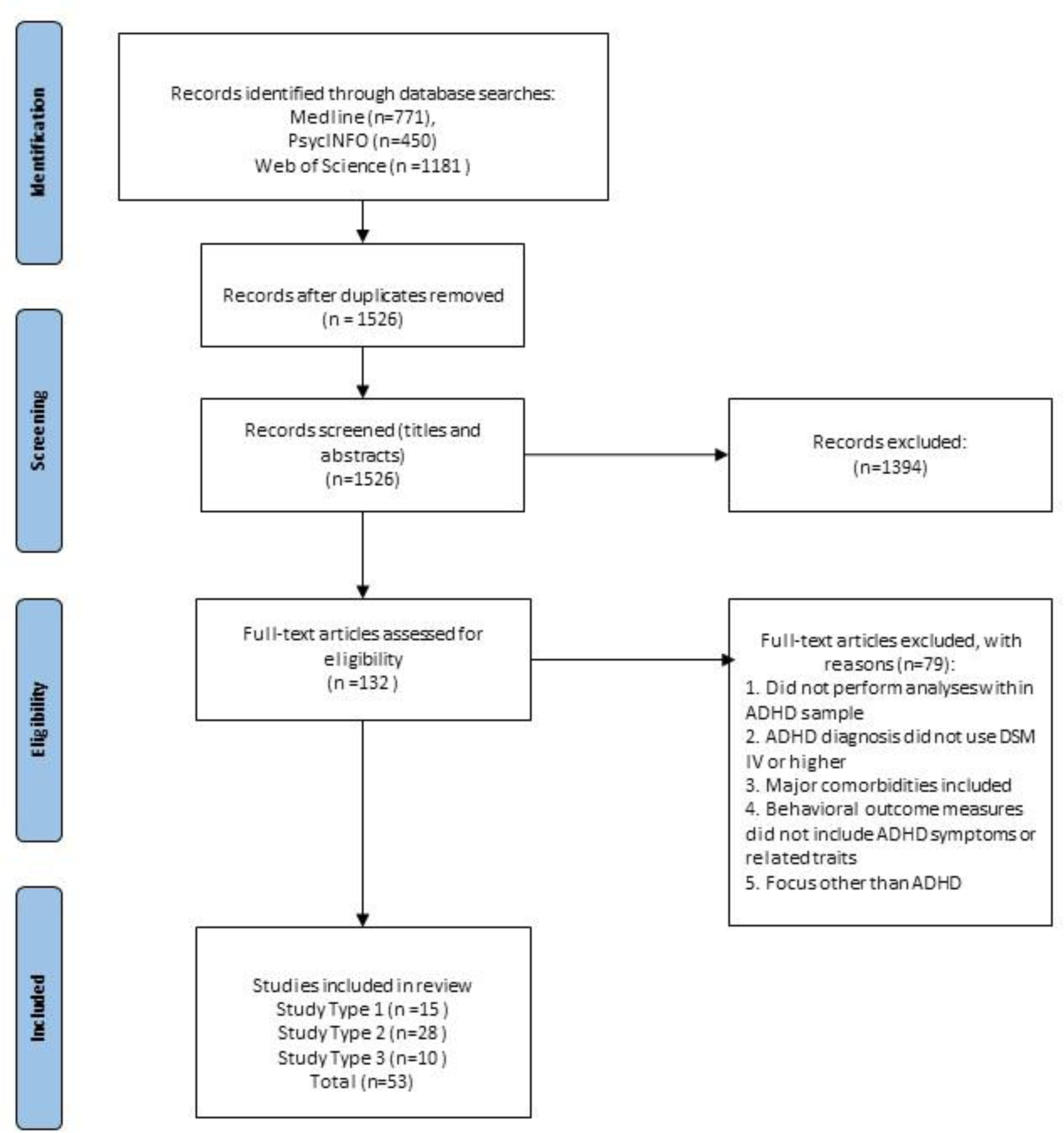
Detailed process of article selection. The flow diagram shows the detailed process of article selection in compliance with the PRISMA guidelines for systematic reviews (Page et al., 2021) http://www.prisma-statement.org/

**Figure 2.**
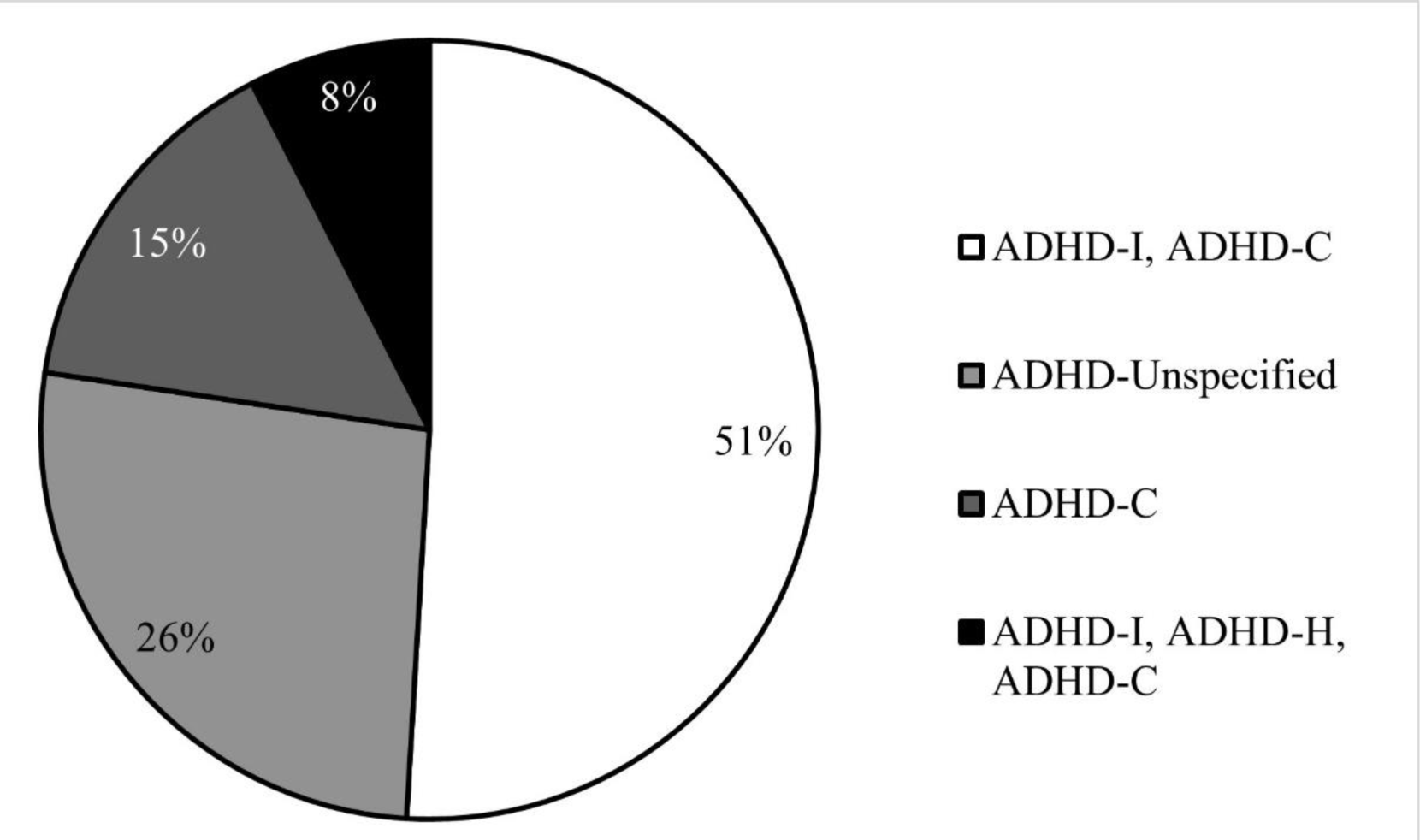
Proportion of included studies by ADHD subtypes examined. The chart summarizes the ADHD subtypes reported in the included studies.

**Figure 3.**
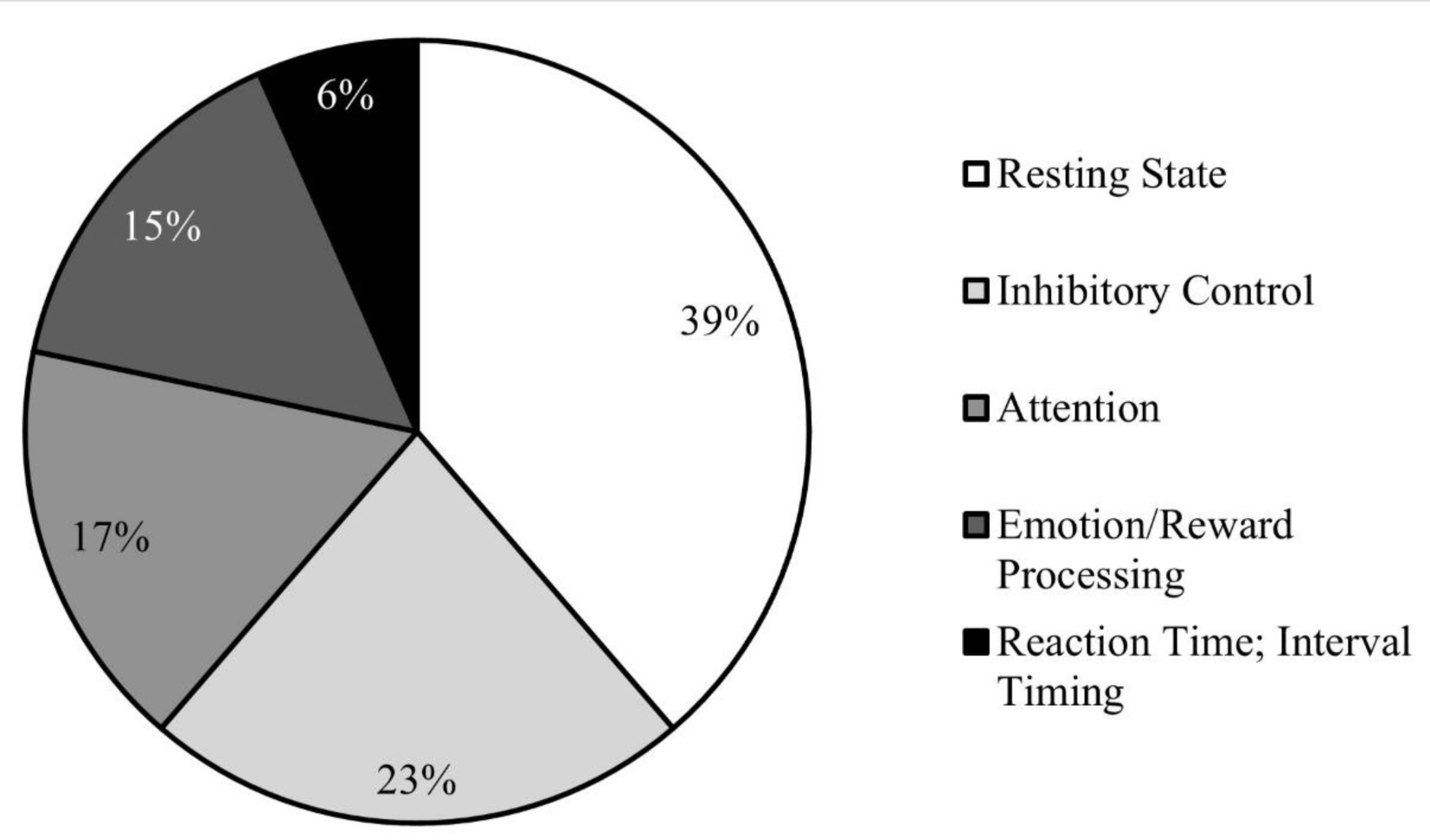
Proportion of included studies by experimental task. The chart summarizes the experimental tasks used during EEG recording in the included studies. Studies that reported data for multiple tasks were included in more than one category accordingly.

In terms of study quality, 27.3% of studies included in the review were rated excellent, 54.7% good, 17% fair, and 0% were rated poor. The median quality score was 16, with a range of 11 to 19. Summary tables are provided for each aim, including extracted data and study quality scores (Tables 1–3).

### 3.1 Are ADHD subtypes associated with different EEG characteristics?

Aim 1 evaluates studies in which EEG characteristics are assessed in two or more of the three diagnostic subtypes of ADHD (predominantly inattentive: ADHD-I; predominantly hyperactive/impulsive: ADHD-H, and combined: ADHD-C). The studies are grouped in this section according to the primary EEG outcome measures used. First, studies evaluating EEG characteristics during resting state tasks (while the individual is not performing an experimental task), using spectral analysis within distinct frequency bands, are evaluated in section 3.1.2. Studies in which EEG was recorded during an experimental task, using either spectral power analysis (section 3.1.3) or event-related potentials (ERPs) are reviewed in section 3.1.4. The studies addressing Aim 1 are summarized and listed alphabetically in Table 1.

#### 3.1.1 General characteristics of the studies

A total of 15 articles met inclusion criteria for Aim 1 (Table 1). Participants’ ages ranged between 7 and 50 years across studies; only one study included adults (Gorman Bozorgpour et al., 2013). The number of ADHD participants included in the studies varied between n=12 and n=164.

#### 3.1.2 Resting state EEG: Spectral analyses

Five of the 15 studies compared resting state EEG between ADHD subtypes. Four of these studies included two subtypes (ADHD-C and ADHD-I) and controls; one study included all three ADHD subtypes and controls. All five studies were with children in the age range of 6-12 years. Two studies included only girls, one study compared subtypes separately in boys and girls, and the remaining two studies included mixed-sex groups. Three of the five studies reported mean spectral power values as well as derived ratio measures, one reported only the beta/theta ratio (the inverse of theta/beta ratio, considered a measure of “cortical activation”) and one used graph theoretical analyses to investigate resting functional network connectivity characteristics associated with beta and theta activity.

The three studies that evaluated mean spectral power all reported elevated slow wave activity (theta and/or delta frequency band) in the ADHD participants compared with controls (Dupuy et al., 2013; Dupuy et al., 2011, 2014), consistent with previous research (Clarke et al., 2001c). Although Dupuy et al. (2011) found no global power differences between the two ADHD subtypes (ADHD-C and ADHD-I) in their girls-only study, differences emerged between subtypes with respect to the topographical distribution of EEG effects across the scalp: compared with ADHD-I, girls with ADHD-C showed higher absolute theta power in the right hemisphere, increased midline posterior beta, reduced right hemisphere relative delta and increased left hemisphere relative theta, reduced midline posterior relative alpha and reduced central relative beta activity (Dupuy et al., 2011). A follow-up study by the same authors recruited girls with a wider range of hyperactive-impulsive symptoms in an attempt to exaggerate potential group differences (Dupuy et al., 2014). As before, no global power differences were observed between the two subtypes, but differences emerged with respect to topographical distribution: girls with ADHD-C demonstrated elevations in midline total power, temporal lobe alpha, and posterior lobe beta, as well as reduced right-hemisphere relative delta, reduced left hemisphere relative alpha activity, and reduced theta/beta ratio (TBR) in the left hemisphere compared with girls with ADHD-I (Dupuy et al., 2014). The 2013 study from the same research group that compared subtype differences in boys and girls separately (Dupuy et al., 2013), showed that ADHD-C boys had globally higher absolute and relative theta, elevated TBR and reduced relative alpha compared with ADHD-I boys, consistent with previous findings in mixed-sex groups (Clarke et al., 2001a; Clarke et al., 2001c), whereas no global power differences were observed between subtypes in the girls (Dupuy et al., 2013), consistent with the results from the girls-only studies discussed above. Although sex differences are beyond the scope of this review, these outcomes from Dupuy et al. highlight that the EEG characteristics associated with ADHD subtypes may also differ between the sexes; this should be taken into consideration in the interpretation of mixed-sex studies, which often include a larger proportion of males than females due to the higher prevalence of ADHD in males in the general population.

One study by Gonzalez-Castro et al. (2010) provided evidence for additive TBR effects between the three subtypes (note that in this study, the calculated measure was the beta/theta ratio, therefore a *lower* value was indicative of a deficit). The beta/theta ratio in ADHD-I was typically low at Cz (central) and normal at FP1 (frontal), and was associated with a behavioral profile of high omission errors and increased reaction times. In contrast, the beta/theta ratio in ADHD-H was normal at Cz but decreased at FP1, associated with fewer omission errors than ADHD-I but high commission errors, increased variability and poor overall executive control. In ADHD-C, there was reduced beta/theta ratio in both frontal and central locations, high omissions and commissions, increased reaction times, increased variability and poor overall executive control. In sum, ADHD-I and ADHD-H were associated with different EEG profiles, and these different profiles were combined in ADHD-C (Gonzalez-Castro et al., 2010).

Finally, Ghaderi et al. (2017) contrasted the three ADHD subtypes in terms of functional brain networks, using graph theoretical analyses. ADHD-C showed decreased global network efficiency and impaired segregation (i.e. lower clustering coefficients) across theta, alpha and beta frequency bands at higher thresholds compared with controls; ADHD-I did not differ from controls. Threshold values were not elaborated (Ghaderi et al., 2017). Further investigation of the differences in network characteristics between subtypes remains an important area for future research.

In summary, the one study that evaluated mean spectral power in both girls and boys revealed global differences between ADHD-I and ADHD-C in the boys only (Dupuy et al., 2013). The other two studies that reported spectral power only in girls reported no global differences between ADHD subtypes but observed differences in the topographical distribution of effects across the scalp among the subtypes (Dupuy et al., 2011, 2014). The single study that included all three ADHD subtypes indicated that atypical beta/theta ratio was observed in different scalp locations in ADHD-I and ADHD-H, and that these atypical features were combined together in ADHD-C (Gonzalez-Castro et al., 2010). Taken together, these findings emphasize the need for additional studies investigating EEG differences between ADHD subtypes that include all three subtypes and take potential sex differences into account.

#### 3.1.3 Task-related EEG: Spectral analyses

Two of the 15 studies addressing Aim 1 used spectral power analysis to investigate task-related EEG activity. One study included children aged 7-14 (Sanchez-Gonzalez and Garcia-Zapirain, 2017) and the other included adolescents, aged 12-17 (Mazaheri et al., 2014). Both studies included mixed-sex groups.

The adolescent study examined ADHD-I, ADHD-C and control participants who performed a cued flanker task (Mazaheri et al., 2014). The suppression of occipital alpha band activity following the presentation of a directional cue is associated with visual processing of the cue, and was significantly affected by group: controls showed the greatest alpha suppression, ADHD-I showed the least, differing significantly from controls; ADHD-C lay in between, and did not differ significantly from ADHD-I or controls. A complementary pattern emerged with regard to the suppression of beta activity contralateral to the cued response hand, which is associated with motor preparation: controls showed the greatest beta suppression, ADHD-C showed the least, differing significantly from controls; ADHD-I lay in between and did not differ significantly from ADHD-C or controls. This study also evaluated trial-by-trial cross-frequency coupling between frontal theta and occipital alpha and reported a significant anti-correlation between post-cue theta and alpha in controls, which was not present in either ADHD subtype (Mazaheri et al., 2014).

The study with younger children investigated suppression of mu rhythm (comparable frequency range to alpha band but associated with sensorimotor areas) during an observation-execution task, in which participants were asked to observe videos of movements (e.g. a video of a subject moving a chess piece rotationally using their left hand) and then imitate the movements themselves (Sanchez-Gonzalez and Garcia-Zapirain, 2017). The study included all three ADHD subtypes but with small sample sizes (6-7 participants per subtype), and overall the study was scored as of lower quality (Table 1). The authors reported differences in task-related suppression of mu activity in frontal, central and parietal areas in ADHD-C and ADHD-H, but only in frontal and parietal areas in ADHD-I; the differences in ADHD-C and ADHD-H were found in the left hemisphere, whereas differences in ADHD-I affected both hemispheres. However, interpretation of these findings was limited by their summary presentation without details regarding the direction of effects (Sanchez-Gonzalez and Garcia-Zapirain, 2017).

In summary, only two studies have investigated task-related spectral analysis of EEG in ADHD subtypes, and one was limited by lower quality. These studies provide evidence for impaired task-related suppression of faster wave activity (alpha and beta band) in ADHD, with ADHD-I showing the greatest deficits in relation to stimulus processing, and ADHD-C showing the greatest deficits in relation to motor preparation. This is an area that warrants further investigation with larger samples that include all three ADHD subtypes.

#### 3.1.4 Task-related EEG: Event-related potentials

Eight of the 15 studies addressing Aim 1 evaluated event-related potentials (ERP) in subtypes of ADHD; all eight studies included only two subtypes (ADHD-I and ADHD-C). Seven of the eight studies evaluated children in the age range of 6-15 years, and one study evaluated adults (18-50 years) (Gorman Bozorgpour et al., 2013). All eight studies included mixed-sex groups. The following experimental tasks were used: three Go/No-Go tasks, two ANT (attention network test), one gambling task, one time interval estimation task and one visual flanker task. The evaluated ERP components differed by task and included both faster (N2) and slower (P3, Contingent Negative Variation (CNV), Lateralized Readiness Potential (LRP)) components locked to a cue or target stimulus, as well as feedback-related components (Feedback-Related Negativity (FRN), Late Lateralized Potential (LLP)).

The first Go/No-Go study used a cued visual task in ADHD-I, ADHD-C and controls (Johnstone and Clarke, 2009). In this cued paradigm, both cue- and stimulus-locked ERP components were evaluated: the CNV is a slower negative cortical potential that appears after the warning cue and before the subsequent stimulus, and is thought to include an early component modulated by noradrenergic systems and a late component related to motor readiness and controlled by dopaminergic systems (Birbaumer et al., 1990; Rohrbaugh et al., 1986). An atypical CNV was observed in both ADHD-I and ADHD-C, with differences in scalp distribution between the subtypes: the CNV was reduced across the midline in ADHD-I and across central regions in ADHD-C. The N2 component is associated with conflict monitoring and response control, and differences in the stimulus-locked N2 latency were observed between the subtypes: the N2 peaked later in ADHD-C than CTRL regardless of stimulus type, while N2 peaked slightly earlier to Go and later for No-Go stimuli for ADHD-I compared with controls (Johnstone and Clarke, 2009).

The second Go/No-Go study (Inoue et al, 2010) used a version of the Continuous Performance Test (CPT) to compare the N2 in adult ADHD participants’ responses to repeated stimuli across trials with switched stimuli across trials. Although there were no significant group effects on behavior, the controls showed an increased N2 in switch trials whereas the ADHD groups did not; no significant differences in the N2 were observed between ADHD-I and ADHD-C (Inoue et al., 2010).

The third Go/No-Go study examined the stimulus-locked Lateralized Readiness Potential (LRP) in adults with ADHD (Gorman Bozorgpour et al., 2013). The LRP is generated in motor and supplementary motor areas, and occurs during response preparation. The early portion reflects initial preparation, prior to the determination of the appropriate response: if the stimulus is determined to be a target, preparation continues and is reflected in the late portion of the LRP; if the stimulus is determined to be a non-target, the potential dissipates and the response is inhibited. Compared with controls, ADHD-I adults responded more slowly and had smaller early LRP components, whereas ADHD-C adults were less accurate, had more variable response times and had smaller late LRP waves (Gorman Bozorgpour et al., 2013).

Two studies (both from the same research group) used the Attention Network Test (ANT), which involves a cued attention task and was originally designed by Fan et al. (2002) to differentiate stages of attentional processing associated with distinct brain networks (Fan et al., 2002). Both studies evaluated the P3 component, which is associated with target detection and attentional allocation, and observed reduced P3 amplitudes in ADHD children compared with controls (Heinrich et al., 2014; Kratz et al., 2011). Reduced cue-P3 amplitude was observed in both ADHD-C and ADHD-I subtypes, whereas reduced target-P3 amplitude was observed only in ADHD-I (Kratz et al., 2011). The follow-up study also evaluated intra-individual range and variability in P3 amplitude across single trials (Heinrich et al., 2014). The single trial data revealed an increased intra-individual range in P3 amplitudes in ADHD-C. Furthermore, range in P3 amplitudes was positively correlated with upper-theta/lower alpha activity (5.5-10.5 Hz) across all participants, suggesting that the increased P3 range observed in ADHD-C may be due to elevated activity in this frequency band. In contrast, ADHD-I showed a reduced intra-individual range in P3 amplitudes, with minimum values comparable to controls but significantly reduced maximum values, indicating deficient resource allocation once the P3 was triggered (Heinrich et al., 2014).

Gong et al. (2014) assessed reward processing in ADHD-I and ADHD-C during a children’s gambling task, under three feedback conditions: large losses, small losses and gains. The feedback related negativity (FRN) and late positive potential (LPP) were evaluated, reflecting early and late stages of feedback processing. ADHD-C showed larger FRN amplitudes in response to large loss feedback than small loss feedback, similar to CTRL, whereas this effect was absent in ADHD-I. ADHD-C also had significantly larger LPP amplitudes than ADHD-I, regardless of feedback condition (neither ADHD subtype differed significantly from CTRL in LPP amplitude). These results indicate that feedback processing is impaired in ADHD-I but intact in ADHD-C, with a trend toward increased processing of negative feedback relative to controls in ADHD-C (Gong et al., 2014).

Bluschke and colleagues (2018) compared the CNV and FRN during a time estimation task in ADHD-C, ADHD-I and CTRL children. ADHD-I showed significantly reduced CNV compared with controls, indicating impaired preparatory processing. ADHD-C showed a slight reduction in CNV amplitudes and significantly reduced FRN compared with controls, indicating impaired feedback mechanisms (Bluschke et al., 2018).

Bluschke and colleagues (2017) compared ADHD-C and ADHD-I children on a visual flanker task (no control group was included in this study). Both groups showed overall effects of stimulus compatibility on behavioral response times, but no differences were observed between the ADHD subtypes in behavior or P3 amplitudes. The authors also used residue iteration decomposition analysis (RIDE) to account for intra-individual variability in the ERP paradigm. Once trial-by-trial variability was taken into account, effects of stimulus compatibility and subtype differences emerged: the RIDE-P3 (analog of the P3) was significantly larger for incompatible trials across all participants; ADHD-C had larger RIDE-P3 overall and showed a greater effect of compatibility on the RIDE-P3 than ADHD-I, driven by significantly larger RIDE-P3 amplitudes in compatible trials (Bluschke et al., 2017).

In summary, several ERP components were atypical in ADHD compared with controls and also showed nuances between the ADHD subtypes. The three Go/NoGo studies revealed impaired response preparation in ADHD compared with controls, reflected in atypical CNV, N2 and LRP components; differences between ADHD subtypes were observed in the scalp distribution of the CNV, the latency of the N2 across stimulus conditions, and the latency and amplitude of early versus late portions of the LRP. The two ANT studies showed deficient attentional allocation, reflected in reduced average P3 amplitudes in both ADHD-C and ADHD-I. The subtypes differed in relation to the potential causes underlying the reduced P3: in ADHD-C, greater range in P3 amplitudes across trials was associated with greater trial-by-trial fluctuations in neural state (in the upper theta/lower alpha frequency band), whereas in ADHD-I, lower maximum P3 amplitudes and reduced range across trials indicated poor resource allocation. In the reward processing task, ADHD-C showed intact feedback processing, with an increased FRN to negative feedback; ADHD-I showed impaired feedback processing, reflected in reduced FRN and LPP components. In the time estimation task it was ADHD-C that showed a reduced FRN, whereas ADHD-I showed a reduced CNV, indicating impaired preparatory processing. Finally, Bluschke et al. (2017) reported that subtype differences in the P3 emerged only once individual variability was taken into account, highlighting that differences between subtypes may be underestimated or “masked” by increased trial-by-trial variability in individuals with ADHD.

### 3.2 Are EEG measures correlated with ADHD traits and/or symptom severity?

This section addresses the second aim: to review studies in which correlational analyses were performed within an ADHD sample to assess relationships between EEG measures and ADHD symptom severity or related traits. These studies are grouped within the section based on the primary EEG measure used, beginning with resting state EEG with spectral analysis in section 3.2.2, task-related EEG with spectral analysis in 3.2.3 and task-related ERP in 3.2.4. It is important to note that the included ERP studies were limited to those in which ERP components were correlated with independent measures of ADHD symptoms or traits, or in which the experimental task could itself be considered a measure of an ADHD-related behavior (e.g. CPT); studies in which ERP components were related to performance on an experimental task, without any independent measure of ADHD symptomatology, were excluded. The studies addressing Aim 2 are summarized and listed alphabetically in Table 2.

#### 3.2.1 General characteristics of the studies

A total of 28 articles met inclusion criteria for Aim 2. Participants’ ages ranged between 6 and 55 years, including 13 studies with children and 15 with adults. The number of ADHD participants included in the studies varied between n=9 and n=93.

#### 3.2.2 Correlations between resting state EEG (spectral analysis) and ADHD symptoms/traits

Seven of the 28 studies assessed correlations between spectral analyses of resting state EEG and ADHD symptoms or traits. Six of the seven studies focused on children within the age range of 7-16 years; one study was with adults (Koehler et al., 2009). Four studies included two ADHD subtypes (ADHD-I and ADHD-C), one study included only ADHD-C participants and two studies included participants with any ADHD diagnosis (number of participants per subtype was not reported). One study included male participants only and the remaining four studies included mixed-sex groups.

All seven studies assessed correlations between resting state EEG characteristics and the core ADHD symptoms of inattention and hyperactivity/impulsivity. In children with ADHD, inattentive symptoms were correlated with elevated global theta power in a sample of boys (Roh et al., 2015), and mixed-sex samples showed correlations of inattentive symptoms with elevated central theta (Ogrim et al., 2012), increased frontal TBR (Zhang et al., 2019), reduced gamma (Barry et al., 2010), and reduced left-lateralized coherence in delta, alpha, beta and gamma bands (Barry et al., 2011). In ADHD adults, inattentive symptoms were correlated with elevated posterior theta (Koehler et al., 2009).

Fewer relationships were reported between resting state EEG and hyperactive/impulsive symptoms. A correlation between hyperactive/impulsive symptoms and lower theta at Cz was reported in children with ADHD (in contrast to the positive correlation between theta and inattentive symptoms reported in the same study, noted above) (Ogrim et al., 2012). A study with boys showed a positive correlation of hyperactive/impulsive symptoms with increased theta (Vollebregt et al., 2015). Vollebregt et al. (2015) also controlled for variation in the TBR due to individual peak alpha frequency, which strengthened the relationship of the TBR with hyperactive/impulsive symptoms (Vollebregt et al., 2015). In the Barry et al. (2011) study described above, hyperactivity/impulsivity was correlated with greater interhemispheric frontal alpha coherence (Barry et al., 2011). The single adult ADHD study reported no significant correlations of EEG spectral analyses of resting state with hyperactive/impulsive symptoms (Koehler et al., 2009).

In summary, inattentive symptoms were correlated with a range of spectral characteristics including elevated theta (in children and adults) and TBR, reduced gamma and reduced left-lateralized coherence across multiple frequency bands. Interestingly, the differences in EEG coherence observed by Barry et al. (2011) provide examples in which greater deviations from the typical EEG profile were associated with fewer ADHD symptoms, suggesting that increased connectivity across a wide frequency range in the right hemisphere may compensate for reduced specialized connectivity in the left hemisphere (Barry et al., 2011). Relationships with hyperactivity/impulsivity were more limited and sometimes inconsistent, possibly due in part to the ADHD samples comprising only ADHD-C and some cases ADHD-I (but no ADHD-H), such that hyperactive/impulsive symptoms were only evaluated in patients who also exhibited inattentive symptoms. Controlling the TBR for individual peak alpha frequency strengthened the correlation between the TBR and hyperactive/impulsive symptoms. Reduced alpha frequency is associated with cortical hypoarousal, whereas elevated TBR is interpreted as an index of delayed cortical maturation; both mechanisms have been proposed as contributing to ADHD but they likely affect EEG characteristics and relate to symptom dimensions in different ways (Clarke et al., 2013).

#### 3.2.3 Correlations between task-related EEG (spectral analysis) and ADHD symptoms/traits

Four of the 28 studies assessed correlations between ADHD symptoms and spectral characteristics of task-related EEG. Two studies included children aged 7-14 years, one included adolescents aged 13-16 years (Helps et al., 2010) and the fourth was with adults aged 18-55 years (Buyck and Wiersema, 2015). One study included two ADHD subtypes (ADHD-I and ADHD-C), one study included only ADHD-C participants and two studies included participants with any ADHD diagnosis (number of participants per subtype was not reported). One study included all males, the rest included mixed-sex groups.

The first study with children recorded EEG while the ADHD and control participants performed a cued visuospatial covert attention task (Guo et al., 2019). Abnormal patterns of alpha modulation were identified during the task in the children with ADHD, and reduced attenuation of alpha activity in the left hemisphere was correlated with inattentive symptoms in the ADHD children but not controls (Guo et al., 2019). The second study with children showed greater frontal alpha asymmetry during a Go/No-Go task that was correlated with commission errors in the ADHD group but not in controls (Ellis et al., 2017). Neither of these studies reported the number of ADHD participants by subtype.

A study with ADHD-C adolescent boys assessed the attenuation of very low frequency activity (02-.2Hz) during the transition from rest to a 2-choice reaction time task (Helps et al., 2010). Persistent low frequency activity during task performance has been posited as indicating intrusion by the default mode network in task-relevant processes (Sonuga-Barke and Castellanos, 2007). Helps et al. (2010) reported that the ADHD adolescents exhibited less rest-task attenuation of low frequency activity than controls, and that lower rest-task attenuation was correlated with higher inattention scores in the ADHD group but not in controls (Helps et al., 2010).

Finally, the adult study that included ADHD-C, ADHD-I and controls recorded EEG during two tasks: a less demanding (slow-paced) 2-choice reaction time task and a more demanding (mid-paced) flanker task. In the slower reaction time task, the ADHD adults performed more slowly and variably, with more commission errors, and also showed increased midline theta and beta power; in the flanker task, ADHD adults showed a comparable effect of congruency to controls (i.e. faster reaction times to congruent versus incongruent stimuli) and no EEG differences were observed compared with controls, indicating intact inhibitory processing (Buyck and Wiersema, 2015).

In summary, two studies showed that inattentive symptoms were associated with impaired suppression of task-irrelevant activity during attentionally demanding tasks. One study reported that commission errors (reflecting impulsivity) were correlated with abnormal alpha lateralization; Buyck and Wiersema (2015) reported increased commission errors associated with increased midline theta and beta activity during a less demanding task. These findings are consistent with evidence for impaired modulation of oscillatory activity during task performance in ADHD (Lenartowicz et al., 2018), although the small number of studies makes it difficult to identify clear patterns in the specific symptom dimensions. The Buyck and Wiersema (2015) study also highlights that in some cases, poor task performance in ADHD may be due to reduced activation during a less demanding task, rather than impaired inhibitory processing per se.

#### 3.2.4 Correlations between task-related EEG (Event-Related Potentials) and ADHD symptoms/traits

Seventeen of the 28 studies investigated correlations between ERP and ADHD symptoms or traits. Four of the 17 studies were conducted with children in the age range of 6-18 years, and 13 studies evaluated adults in the age range of 18-51 years. All ADHD participants included in the studies were ADHD-C, ADHD-I or of unspecified subtype, except for one study which included all three subtypes. Two studies included male participants only; the rest included mixed-sex groups. The following experimental tasks were used: six Go/No-Go, five emotion and reward processing, three auditory oddball, one ANT (Attention Network Test), one visual search and one CPT (Continuous Performance Test). The ERP components differed by task and included a range of faster (N1; N170; N2; ERN) and slower (CVN; LPP; Pe; P3, RPNS) stimulus-locked components, as well as the feedback-related negativity (FRN).

Woltering et al. (2013) administered a visual Go/No-Go task to adults with and without ADHD (subtypes were not specified). Smaller N2 amplitudes were correlated with poorer performance on the No-Go trials only in the ADHD group, and reduced P3 amplitudes were associated with inattentive symptoms but not with hyperactive/impulsive symptoms (Woltering et al., 2013). Grane et al. (2016) also used a visual Go/No-Go paradigm with ADHD-C adults and reported that reduced P3 amplitudes to both Go and No-Go stimuli were associated with greater response variability and more omission errors (Grane et al., 2016). Kakuszi et al. (2016) recorded EEG in ADHD-C adults performing a Go/No-Go task, and also had participants complete a separate Stroop task. The Response Preceding Negative Shift (RPNS) in fronto-central brain regions was evaluated as a measure of response preparation. Adults with ADHD showed an increased RPNS in correct Go trials compared with controls; larger amplitude RPNS was correlated with poorer performance on the Stroop task, increased response variability and increased hyperactive/impulsive symptom severity (Kakuszi et al., 2016).

Another Go/No-Go study with ADHD-C adults and controls assessed the error-related negativity (ERN) and error positivity (Pe) components (Czobor et al., 2017). Reduced amplitudes in both components were observed in the adults with ADHD compared with controls, and different scalp distributions of these potentials were correlated with different symptom dimensions within the ADHD group only: increased hyperactivity was associated with reduced ERN amplitudes in right temporal and lateral parietal areas, and increased impulsivity was correlated with reduced Pe amplitudes in middle frontal regions. Increased inattentive symptoms were associated with reduced ERN in right lateral parietal areas, and reduced Pe in posterior-parietal areas (Czobor et al., 2017).

Seven studies investigated the relationship of ADHD symptoms to ERP components during emotion and reward processing tasks. A study with ADHD adolescents included all three subtypes (although only two participants were ADHD-H) and used a three-condition emotional Go/No-Go task to investigate the N170, which is associated with facial emotion processing (Alperin et al., 2017). Larger N170 amplitudes in the happy condition were related to higher parental rankings of positive/approach emotions in the ADHD children (Alperin et al., 2017). Ibanez et al (2011) examined adults with ADHD performing a facial emotion processing task; reduced N170 amplitudes were correlated with poorer performance on an emotional inference Theory of Mind task, and reduced N170 amplitudes in response to simultaneous stimuli were related to poorer executive function in the ADHD sample (Ibanez et al., 2011). Another study used an emotional version of the Go/No-Go task with adult males, reporting that the ADHD adults showed a reduced right parietal late positivity (LPP) when instructed to inhibit a response to negative emotions (Kochel et al, 2012). Although there were no objective behavioral differences between the ADHD and control groups, this reduced positivity was correlated with lower self-reported emotional intelligence within the ADHD group. ADHD subtypes were not specified in this study (Kochel et al., 2012).

Shushakova et al. (2018a) administered an emotional regulation task to ADHD-C and ADHD-I adults. They reported that increased frontal and centroparietal late positive potential (LPP) amplitudes were correlated with poorer self-reported emotional regulation skills and higher ADHD symptom severity (inattention and hyperactivity/impulsivity were not reported separately) (Shushakova et al., 2018a). A study investigating attentional bias in adults with ADHD revealed that the normal bias to positive words, found in healthy controls, was not present in ADHD (Shushakova et al., 2018b). Instead, individuals with ADHD showed a bias toward both positive and negative emotional words compared with neutral words, reflected in increased N2pc amplitudes. These increased amplitudes were correlated with poorer task performance and increased ADHD symptoms (Shushakova et al., 2018b). Both studies from Shushakova et al. included ADHD-C and ADHD-I participants.

Cheung et al. (2017) administered a four-choice reaction time task to young adults with ADHD-C adults and a control group. Reduced P3 amplitudes in the ADHD group were correlated with higher reaction time variability, and a greater increase in P3 amplitudes from baseline to the fast-paced rewarded condition was correlated with a decrease in response variability. In contrast with controls, the ADHD group did not show an increase in CNV amplitude between baseline and reward conditions (Cheung et al., 2017). Thoma et al. (2015) assessed the FRN during a probabilistic reward-based learning task in adults with ADHD-C and controls. No significant correlations were observed between ADHD symptom severity and FRN amplitudes (Thoma et al., 2015).

Three studies used an auditory oddball task to assess attentional processing and sensory gating in ADHD. Increased ADHD symptom severity in a child sample was correlated with reduced P3b amplitudes in response to auditory targets (Janssen et al., 2016) and with reduced MMN amplitudes to deviant tones at Cz and CPz (Lee et al., 2020). Micoulaud-Franchi et al. (2019) reported that lower P300 amplitudes in an adult ADHD sample were correlated with higher self-reports of distractibility but not with clinical symptom severity (Micoulaud-Franchi et al., 2019).

Abramov et al. (2019) used the attention network test (ANT) to investigate attention-related ERP components in boys with and without ADHD; reduced P3 amplitudes were observed in the ADHD sample, but were not correlated with symptom scores. Interestingly, increased rightward lateralization in occipital areas 40-200 ms after target onset was observed in the boys with ADHD, but greater occipital asymmetry (i.e. increased deviation from the typical pattern) was correlated with fewer inattentive symptoms in the ADHD group, potentially reflecting compensatory right hemisphere mechanisms in children with ADHD (Abramov et al., 2019).

Cross-Villasana et al (2015) studied an adult ADHD sample and a control sample performing a visual search paradigm. Increased delays in posterior contralateral negativity (PCN) components were correlated with increased symptom severity in the ADHD group, reflecting slowed attentional selection (correlations were not reported in the control group) (Cross-Villasana et al., 2015).

Kaur et al (2019) studied an ADHD adult sample and a control sample performing a visual continuous performance attention task. Increased latency of the N2 in response to standard stimuli was positively correlated with symptom severity. Smaller P3 and N2 amplitudes and longer N1 latencies in response to the target stimulus were correlated with increased symptom scores. This study also evaluated additional characteristics of the EEG signal including entropy, and determined that reduced entropy in the theta range was correlated with higher symptom scores. (Total ADHD symptom scores were calculated for the ADHD participants only, and subscale scores were not computed; correlations with symptom severity were therefore only reported within the ADHD sample) (Kaur et al., 2019).

In summary, several relationships emerged between atypical ERP components observed in ADHD, and specific ADHD symptoms or related traits. Components reflecting impaired response control (reduced N2) and response preparation (increased RPNS) were associated with increased hyperactivity/impulsivity symptoms and commission errors (also a measure of impulsivity). Potentials linked to poor attentional allocation (reduced P3) were associated with increased inattentive symptoms, omission errors (also a measure of inattention), total ADHD symptoms, greater response variability and self-reported distractibility. Potentials relating to emotion and reward processing (N170 and LPP) varied according to context: reduced N170 amplitude was associated with poor performance on an emotional inference task and increased N170 to positive emotions was associated with parental reports of positive/approach behavior, whereas increased N170 to negative emotions was associated with poor emotional regulation and increased LPP amplitudes were associated with increased total ADHD symptoms. Individuals with ADHD also showed a stronger bias toward positive and negative emotional words, reflected in increased N2pc amplitudes that were correlated with poorer task performance and increased total ADHD symptoms.

Similar findings of ERP reduction corresponding to increased ADHD symptoms were reported for the ERN, Pe, and right parietal late positivity, with different scalp distributions of these potentials relating with different symptom dimensions: potentials correlated with hyperactive/impulsive symptoms had more widespread distribution whereas those correlated with inattentive symptoms were constrained to parietal and post-parietal areas. Interestingly, several studies reported increased latencies in ERP components among ADHD samples, including PCN components and N2 components during attentional tasks, that were consistent with longer behavioral processing times, and were correlated with increased ADHD symptom severity. Finally, there was evidence for increased rightward asymmetry of stimulus processing in ADHD, potentially reflecting compensatory mechanisms for less efficient left hemisphere function.

Limitations included reports of total ADHD symptom scores, without the separate subscale scores that would permit evaluation of relationships between ERP components and distinct symptom dimensions (inattention or hyperactivity/impulsivity). Many studies reviewed here included only one or two ADHD subtypes; investigation of relationships between EEG characteristics and ADHD symptom dimensions across all three subtypes is an important area for future research.

### 3.3 Do classification techniques using EEG measures reveal different clinical presentations of ADHD?

The final section assesses studies in which EEG characteristics were used to identify subgroups in an ADHD sample. Section 3.3.2 reviews studies in which EEG measures were used to classify patients according to diagnostic subtype; section 3.3.3 reviews studies in which patients were grouped based on previously-identified EEG markers (e.g. elevated TBR) and then compared on behavioral or other EEG measures; section 3.3.4 reviews studies in which novel subgroups were discovered based on EEG characteristics. The studies addressing Aim 3 are summarized and listed alphabetically by author in Table 3.

#### 3.3.1 General characteristics of the studies

A total of ten articles met inclusion criteria for Aim 3. Participants’ ages ranged between 5 and 55 years; nine studies included children only (one of which included participants up to 21 years) and one study included both children and adults (7-14 years and 18-55 years) (Buyck and Wiersema, 2014). The number of ADHD participants included in the studies varied between n=30 and n=620. All studies included mixed-sex groups. The classification and grouping methods included cluster analysis, latent class analysis, deep learning and decision tree algorithms and Bayesian Gaussian Mixture models as well as manual grouping based on visual inspection of EEG signals or defined thresholds in variables of interest (spectral power, TBR or ERP amplitude measures). Eight studies collected original data, one study applied analyses to archived clinical data (Martin and Konopka, 2011) and one used data collected in an earlier study (Robbie et al., 2016).

#### 3.3.2 Classification of ADHD diagnostic subtypes using EEG

Three of the ten studies used EEG measures to classify participants based on existing ADHD subtypes. Two studies used spectral analysis of resting state EEG and one study used single-trial EEG. Two studies included only children and one included both children and adults. Two studies included only two ADHD subtypes (ADHD-I and ADHD-C) and one study included all three subtypes.

The first study investigated the use of single-trial EEG to distinguish between ADHD-C and ADHD-I subtypes in children, using a deep learning approach (Vahid et al., 2019). The experimenters collected EEG data during a time estimation task, in which participants were asked to press a button when 1200 ms had elapsed, and they were given feedback about their response accuracy. Using these EEG data, the classifier was able to distinguish both ADHD subtypes from controls but did not succeed at differentiating the subtypes from one another (Vahid et al., 2019).

The second study applied Receiver Operation Characteristic (ROC) curves to evaluate the effectiveness of regression models in predicting ADHD subtypes (Buyck and Wiersema, 2014). The models used resting state EEG (spectral analyses) in children and adults with ADHD, including both ADHD-I and ADHD-C subtypes. Elevated TBR was observed in both children and adults with ADHD-I, compared with ADHD-C or controls; however, these EEG differences were not sufficient to enable accurate classification of individual participants into diagnostic subtypes (Buyck and Wiersema, 2014).

The third study applied a decision tree algorithm to resting state EEG to classify ADHD subtypes (including all three subtypes, ADHD-I, ADHD-H and ADHD-C) and controls (Rostami et al., 2020). The algorithm used a combination of the Child Behavior Checklist (CBCL), Integrated Visual and Auditory (IVA) test, and spectral power in the delta, theta, alpha and beta bands, as well as TBR. A perfect classification accuracy (100%) was attained for the categories of ADHD versus controls; the algorithm was also able to discriminate among the three ADHD subtypes with an accuracy of 80.41% (ADHD-C), 84.17% (ADHD-I), and 71.46% (ADHD-H), respectively (Rostami et al., 2020).

In summary, the small number of studies attempting to classify existing ADHD subtypes using EEG characteristics had mixed success. The most successful classification outcomes relied upon a combination of neuropsychological measures with resting state EEG to classify all three subtypes; therefore, it remains unknown how successful the classification would be with only EEG measures. It is possible that the inclusion of ADHD-H (which is included in fewer studies overall) enabled clearer differentiation of the EEG characteristics associated with inattentive and hyperactive/impulsive symptom dimensions, resulting in better performance of the classifier.

#### 3.3.3 Investigation of previously-defined EEG subgroups

Five of the ten studies used previously-identified EEG markers to create ADHD subgroups, and then assessed differences in EEG or behavioral measures between the subgroups. Four studies used spectral analysis of resting state EEG and one study defined subgroups based on the error-related negativity (ERN) during a reaction time task (Liao et al., 2018). All five studies included children. One study included only one subtype (ADHD-C), three studies included two ADHD subtypes (ADHD-I and ADHD-C) and one study included all three subtypes.

Zhang et al. (2017) investigated relationships between elevated TBR and inhibitory functions in children with ADHD-I and ADHD-C and a control group. The ADHD sample was divided into two subgroups: one with elevated TBR (top 35% of TBR values, n=20) and one with normal TBR (n=38). The elevated TBR group showed greater difficulty inhibiting conflicting stimuli in a flanker task (inhibitory control) but reported fewer inhibitory issues on a self-report questionnaire of social inhibition. Performance on a stop-signal task (response inhibition) did not differ between the ADHD subgroups (Zhang et al., 2017).

Martin and Konopka (2011) applied cluster analysis to archival patient data to examine three previously defined clusters of abnormal resting state EEG (Chabot and Serfontein, 1996; Clarke et al., 2001b; Monastra et al., 2001) in ADHD-C and ADHD-I patients. The three clusters comprised: a “cortical hypoarousal” cluster characterized by excess theta, a “cortical hyperarousal” cluster with excess beta and decreased theta/delta, and a “maturation lagged” cluster with excess delta and theta. The authors noted an unexpectedly high proportion of cortically hyperaroused children, i.e. exhibiting elevated beta activity, which may have resulted from the larger proportion of ADHD-C in the group (26 ADHD-C and four ADHD-I). In addition, the authors identified a novel “combined” subgroup, characterized by elevations in both parietal beta and central theta frequencies (Martin and Konopka, 2011).

Robbie et al. (2016) used data collected in a previous study (Clarke et al., 2011), including two of the five EEG-based ADHD subgroups identified via cluster analysis in that study and age-matched controls. The ADHD subgroups had either excess theta or excess alpha power present in their resting state EEG. Interhemispheric and intrahemispheric EEG coherence (temporal correlation between two signals in a frequency band) was calculated within the delta, theta, alpha and beta frequency bands. Compared with controls, both ADHD subgroups had increased theta intrahemispheric coherence at shorter inter-electrode distances. The excess-theta subgroup also had increased frontal interhemispheric theta and reduced beta coherences. The excess-alpha subgroup showed reduced longer-range coherence in the alpha band (Robbie et al., 2016).

Another study used visual analysis to group ADHD children and controls based on three pre-determined patterns of atypical resting state EEG (Machinskaya et al., 2014). The three EEG-defined patterns included one subgroup exhibiting atypical fronto-thalamic functioning (bilaterally synchronous frontal theta waves, or FTW), a second subgroup with local deviations (sharp theta waves) in the right hemisphere, and a third group with deficits in nonspecific activation, such as hypersynchronous alpha or slow-wave activity in caudal areas. Compared with controls, the children in the abnormal FTW group showed significant difficulties with executive function and verbal tasks, whereas children with right hemisphere theta wave differences showed greater non-verbal deficits in visual perception and visual memory (Machinskaya et al., 2014).

Liao et al. (2018) grouped children with ADHD-I or ADHD-C based on their behavioral errors and corresponding error-related negativity (ERN) component during a simple 4-minute reaction time task. Two novel ADHD groups were created: one with reduced ERN amplitudes (n=26) and one with ERN amplitudes comparable to controls (n=5). Only the reduced-ERN subgroup had poorer performance on the Comprehensive Nonverbal Attention test, an objective measure of executive function, although both groups provided equivalent subjective ratings of executive function (Liao et al., 2018).

In summary, these studies support the presence of distinct ADHD subgroups based on previously-identified EEG markers, including spectral analyses of resting state EEG or from ERP components such as ERN during executive function tasks. Furthermore, groups of individuals with more extreme TBR or frontal theta showed some correspondence to decreased inhibitory function. Although some differences in EEG coherence were found in the excess-theta and excess-beta subgroups, similar findings of increased slow wave activity across the ADHD subgroups led the authors to argue that frontal theta networks were disrupted in the majority of ADHD cases. However, more research is needed to determine which EEG characteristics are most reliable across samples and to determine clearer associations between EEG and symptom profiles.

#### 3.3.4 EEG-based discovery of novel ADHD subgroups

Two of the ten studies used EEG characteristics to discover novel ADHD subgroups. Both studies used spectral analysis of resting state EEG, and both included children and adolescents (ages ranged from 5 to 21 years). One study included ADHD participants of any subtype and the other included ADHD-I and ADHD-C, but neither reported the number of participants per subtype.

Bussalb et al. (2019) evaluated resting state EEG in a large sample of ADHD-C and ADHD-I (n=363), aged 5-21 years, and applied Bayesian Gaussian Mixture models to define novel patient subgroups based on spectral EEG characteristics. Two distinct components were identified, one of which had normal TBR, and one of which had elevated TBR (comprising 36% of the sample). Behavioral measures were not compared between the subgroups (Bussalb et al., 2019).

Loo et al. (2018) evaluated resting state EEG in a large sample of children with and without ADHD; all subtypes were included (number of participants in each subtype were not reported). Using latent class analysis of the EEG signal, the researchers determined that a five-class model provided the best fit overall, with the ADHD and control children distributed across the five classes. Four of the five EEG-defined classes were characterized by spectral power elevations in a specific frequency band (delta, theta, alpha or beta), whereas the fifth class showed no spectral elevation. The clusters with elevated slow-wave activity (classes defined by elevated delta or theta activity) demonstrated higher levels of externalizing behaviors and cognitive deficits, whereas the classes with elevated alpha and beta power had higher levels of internalizing behaviors and emotion dysregulation, with intact cognitive functioning. However, the distribution of ADHD and control children was similar across the classes, indicating that there is not a distinct resting EEG profile associated with ADHD diagnosis. The EEG-based subgroups differed in mean age, with the youngest children in the elevated theta subgroup and the oldest children in the elevated beta subgroup (Loo et al., 2018). This pattern is consistent with an increase in faster (higher frequency) EEG wavelength activity (shift from theta to beta) during development (Marek et al., 2018).

In summary, these studies support the presence of distinct EEG-based clusters of individuals in ADHD samples, that can be identified based on spectral analyses of resting state EEG. It is important to note that in the Loo et al. (2018) study, the clusters observed within the ADHD samples were also observed in control participants: this emphasizes the importance of a multi-dimensional approach to symptoms, as ADHD group effects may be qualified by similar heterogeneity within the control population. Although this section represents only two studies, the increasing sophistication of available computational methods offers great promise for future research in this area.

## 4. Discussion

The central objective of this review was to investigate variation in EEG characteristics in children and adults diagnosed with ADHD, a broad diagnostic category that includes three clinical subtypes (predominantly inattentive; predominantly hyperactive; combined inattentive / hyperactive). To our knowledge, this is the first systematic review to focus on individual differences in EEG characteristics in subtypes of the ADHD population, and to assess both spectral power characteristics and event-related potentials in a range of tasks as well as in resting state. Figure 2 displays the primary focus of most studies has been to compare the inattentive and combined subtypes, and highlights the paucity of studies that include all ADHD subtypes. Figure 3 demonstrates the research focus on inhibitory control tasks and attention tasks, as well as resting state EEG; these tasks tie most directly to the symptomatology that distinguishes the ADHD subtypes.

Three distinct aims were addressed that integrated the EEG findings across study types. Aim 1 focused on differences in EEG measures between existing ADHD subtypes. Aim 2 focused on correspondences between EEG markers and ADHD symptomatology. Aim 3 addressed classification methods of EEG signals that reveal novel ADHD subgroups. Of the 53 studies included in the review, 37 included children and 17 included adults (one study included both). The majority of adult studies (15 of 17) was found in Aim 2. A significantly larger proportion of the ERP studies (17 of 26) was also found in Aim 2. Differences in the distribution of studies across the three aims may reflect a more clinical focus or goal of identifying diagnostic markers (particularly in children) to identify (Aim 1) or to classify (Aim 3) ADHD subtypes, compared with the experimental focus of the correlational studies (Aim 2).

### Aim 1: Differences in EEG measures between ADHD subtypes

These studies expanded upon previous findings in group comparisons of ADHD and controls, revealing nuances in EEG markers between the subtypes. Elevated slow wave activity (theta and/or delta frequency band) was consistently observed in ADHD participants, with differences emerging between the subtypes in the topographical distribution of the elevated activity across the scalp. Impaired task-related suppression of faster wave activity (alpha and beta band) was observed in ADHD, with ADHD-I showing deficits in stimulus processing, and ADHD-C showing deficits in motor response preparation.

The ERP studies also showed subtle differences between the ADHD subtypes, including differences in the topographical distribution of ERP components across the scalp. In one study, subtype differences in the P3 emerged only once individual variability was taken into account, highlighting that differences in averaged ERP components may be masked by increased trial-by-trial variability in individuals with ADHD. Outcomes related to feedback and reward processing were more task-dependent, likely reflecting the interaction between energetic or motivational state and cognitive processing in these tasks.

### Aim 2: Correlational patterns between EEG measures and ADHD symptom severity or related traits

Inattentive symptoms were correlated with a range of spectral characteristics, including elevated theta (in children and adults) and TBR, reduced gamma and reduced left-lateralized coherence across multiple frequency bands. Inattentive symptoms were also associated with impaired suppression of task-irrelevant activity (i.e. alpha-band and very low frequency activity) during attentionally demanding tasks. Fewer relationships with hyperactivity/impulsivity were observed, with conflicting outcomes in relationships to theta power across studies. In one study, controlling for individual peak alpha frequency strengthened the correlation between hyperactive/impulsive symptoms and the TBR, again suggesting differences between subtypes may be masked by other factors (in this case cortical hypoarousal). ADHD-related effects on task performance and task-related EEG activity were also influenced by task difficulty, potentially reflecting an interaction with reduced overall activation during less demanding tasks. Interestingly, in one study, greater deviations from the typical EEG profile were associated with fewer ADHD symptoms and suggested potential right hemisphere compensatory mechanisms.

The ERP studies in Aim 2 provided wide-ranging evidence for relationships between ERP components and specific ADHD symptoms or related traits, although there was only a small number of studies assessing each measure (see Figure 2). As expected, hyperactivity and impulsivity were associated with neural indices of disrupted response control and response preparation whereas inattention was associated with components reflecting disrupted attentional allocation. Poor emotional regulation was associated with increased neural responses to emotion, although increased neural responses specifically to positive facial emotion were also linked to increased positive/approach behavior; impaired emotional inference was associated with reduced neural processing of emotion. These findings emphasize that ADHD-related deficits often reflect an inappropriate allocation or poor regulation of neural resources, rather than a simple increase or decrease in a given component.

### Aim 3: Classification measures of EEG revealing different clinical presentations of ADHD

The studies attempting to classify existing ADHD subtypes using EEG characteristics had mixed success, with the most successful outcomes relying on a decision tree classification algorithm, and using a combination of neuropsychological and electrophysiological measures. Several studies used previously-identified EEG markers to create ADHD subgroups and demonstrated that subgroups with elevated TBR or frontal theta exhibited poorer inhibitory function. The two studies using EEG measures to discover novel EEG-based clusters in the ADHD sample revealed different clusters; more studies are needed to determine whether EEG-based subgroups can be consistently identified across ADHD samples and linked to distinct behavioral profiles. It is important to note that in one study the clusters observed within the ADHD samples were also observed in control participants: this emphasizes the importance of a multi-dimensional approach to symptoms, as ADHD group effects may be qualified by similar heterogeneity within the control population.

### 4.1 Limitations

The majority of studies reviewed here included only one or two ADHD subtypes (as is reflected in the summary of studies by included subtypes in Figure 2), most often the predominantly inattentive and combined subtypes. The presence of both inattentive and hyperactive/impulsive symptoms in the combined subtype make it more difficult to disentangle specific EEG markers associated with each symptom dimension when only the inattentive and combined subtypes are compared. Those studies that included all three subtypes provided some evidence for distinct EEG profiles in ADHD-I and ADHD-H, with additive effects observed in ADHD-C. Due to the wide range of experimental tasks and measures used across the studies, several sub-sections contained very few studies. Often, subtype or correlational analyses were included as secondary analyses in a study that was primarily focused on comparisons between ADHD and controls. This review highlights the need for studies that are focused on the investigation of individual differences in EEG characteristics and related behavior in ADHD.

### 4.2 Conclusions and future directions

The studies in this review provide strong support for the use of electrophysiological measures to provide meaningful insights into the heterogeneity of ADHD. Although the studies presented here do not yet support direct clinical translation of EEG biomarkers for diagnostic purposes, continued research that emphasizes investigation of individual differences rather than a “one size fits all” approach to ADHD has the potential to inform and refine the clinical management of ADHD.

Future research directions should include investigation of the relationship between atypical spectral power profiles and disrupted ERP components in individuals with ADHD. In particular, elevated slow wave activity may contribute to increased trial-by-trial variability in neural responses, which in turn may have a significant impact on averaged event-related components. Although some interesting relationships emerged with the existing diagnostic symptom categories of inattention and hyperactivity/impulsivity, global cognitive functions such as attention rely on highly distributed neural processing, and symptoms of inattention may therefore result from multiple underlying causes. It is likely that behavioral tasks more closely tied to specific underlying brain mechanisms (e.g. error processing, timing behavior, reward sensitivity) may reveal stronger relationships with electrophysiology. Key measures that show promise for the discrimination of existing ADHD subtypes and symptom dimensions include: resting state alpha, beta and theta power, task-related modulation (both suppression and hemispheric lateralization) of alpha and beta activity, and the event-related N2 and P3 components. Additional measures relating to reward processing, emotional regulation and temporal processing may prove informative for discovering novel EEG-based subgroups, since these are ADHD-related traits not currently reflected in the DSM-5. It is likely that a more nuanced characterization of ADHD subtypes and symptom dimensions will evolve alongside the investigation of EEG-based markers.

Increasing evidence suggests that ADHD is associated with multiple etiological pathways. Future research examining individual differences in EEG characteristics in relation to genetic risk factors and the development of structural and functional brain networks could provide valuable insights into the causal mechanisms underlying ADHD. The availability of large datasets and increasing sophistication of computational methods make the application of classification techniques and machine learning algorithms an especially promising area for future research, both in the discrimination of previously-defined ADHD subtypes and the discovery of novel EEG-based subtypes.

## Declaration of Competing Interest

The authors report no declarations of competing interest.

## Data Availability

All data produced are contained within the manuscript.

## Acknowledgements

This work was supported in part by postdoctoral funding to J. Slater from the Canada First Research Excellence Fund, awarded to McGill University for the Healthy Brains, Healthy Lives initiative. This work was also supported by the Natural Sciences and Engineering Research Council of Canada (grant 298173) and by a Canada Research Chairs grant to C. Palmer. The authors would also like to thank Emily Kingsland for library assistance with the search strategy, database searches and guidance with the systematic review process.

## Notes

### Competing Interest Statement

The authors have declared no competing interest.

### Funding Statement

This study was funded in part by postdoctoral funding to J. Slater from the Canada First Research Excellence Fund, awarded to McGill University for the Healthy Brains, Healthy Lives initiative. This work was also supported by the Natural Sciences and Engineering Research Council of Canada (grant 298173) and by a Canada Research Chairs grant to C. Palmer.

### Author Declarations

The systematic review includes only previously published data.

